# Evaluating scope and bias of population-level measles serosurveys: a systematic review and bias assessment

**DOI:** 10.1101/2023.08.29.23294789

**Authors:** Alyssa N. Sbarra, Felicity T. Cutts, Han Fu, Ishu Poudyal, Dale Rhoda, Jonathan F. Mosser, Mark Jit

## Abstract

**Background:** Measles seroprevalence data has potential to be a useful tool for understanding transmission dynamics and for decision making efforts to strengthen immunization programs. In this study, we conducted a systematic review and bias assessment of all primary data on measles seroprevalence in low- and middle-income countries published from 1962 to 2021.

**Methods:** On March 9, 2022, we searched PubMed for all available data. We included studies containing primary data on measles seroprevalence and excluded studies if they were clinical trials or brief reports, from only health care workers, suspected measles cases, or only vaccinated persons. We extracted all available information on measles seroprevalence, study design, and seroassay protocol. We conducted a bias assessment based on multiple categories and classified each study as having low, moderate, severe, or critical bias. This review was registered with PROSPERO (CRD42022326075).

**Findings:** We identified 221 relevant studies across all World Health Organization regions, decades and unique age ranges. The overall crude mean seroprevalence across all studies was 78.00% (SD: 19.29%) and median seroprevalence was 84.00% (IQR: 72.75 – 91.66%). We classified 80 (36.2%) studies to have severe or critical overall bias. Studies from country-years with lower measles vaccine coverage or higher measles incidence had higher overall bias.

**Interpretation:** While many studies have underlying bias, many studies provide data that can be used to inform modelling efforts to examine measles dynamics and programmatic decisions to reduce measles susceptibility.

**Funding:** Bill & Melinda Gates Foundation; Gavi, the Vaccine Alliance; US National Institutes of Health

**Research in Context:** *Evidence before this study:* On August 20, 2023, we searched PubMed for systematic reviews published from January 1, 1980 to August 20, 2023 using the search terms “measles” AND “sero*”. We included studies if they were a systematic review of measles seroprevalence data and excluded studies that did not contain information on measles seroprevalence, were not systematic reviews, only included data from persons within a subpopulation (e.g., pregnant people or healthcare workers), or were of head-to-head laboratory comparisons of assay methodology. We identified one previous systematic review, by Thompson and Odahowski, published in 2016 and including data through mid-2014. That review identified 220 measles and/or rubella seroprevalence studies from all countries globally. Study authors published a descriptive summary of seroprevalence trends by age in a five select countries and a narrative summary of high-level epidemiologic trends in the underlying data, including information available on maternal antibody waning. Beyond these select summary findings, that study did not separately report seroprevalence from each study identified in the analysis, nor did it include any information on study design or population-representativeness. While study authors noted general limitations related to the different methods used across studies, they did not include any specific information on assay type, selection biases or other characteristics that could influence the accuracy of results or include data in a tabular format, which limits the utility of this study for subsequent analyses.

*Added value of this study:* Our study builds upon the known body of data on measles seroprevalence from low- and middle-income countries in multiple ways. First, we included data published up to December 31, 2021 and from non-English language studies. Second, we extracted all available relevant information on study design characteristics and assay protocol used in each study to measure seroprevalence. Then, we constructed a bias assessment framework and conducted a bias assessment across multiple categories (study selection of participants, measurement tool and classification of immunity, and reporting of results) to classify the underlying bias in each study. Finally, we compared seroprevalence estimates across regions and bias levels, and bias levels among various study location characteristics.

*Implications of all the available evidence:* Accounting for study design and seroassay protocol used in serosurveys can influence interpretation of population-level seroprevalence estimates. Our systematic review and bias assessment provides an updated landscape of serological studies and highlights key biases in the current literature. It provides a repository of measles seroprevalence data, along with corresponding critical information on factors that influence population-representativeness and overall sensitivity of the measurement assay used in each study, that can be used to inform measles susceptibility estimates useful for planning targeted vaccination efforts.

## Introduction

Measles remains a substantial cause of global morbidity and mortality^1^, especially in low- and middle-income settings where over 99% of measles cases and deaths occur^2^, despite the availability of a safe and effective vaccine^3^. Because ongoing measles transmission can be maintained if herd immunity (i.e., when the proportion of the population immune is sufficient to limit disease spread) has not been reached and sustained, estimating the proportion of people susceptible within a community is essential to plan immunization programs and assess future risk of measles outbreaks and deaths. However, due to factors such as timeliness of and age at vaccination^4^, disruptions to cold chains^5^, a lack of seroconversion in specific subpopulations (e.g., among persons living with human immunodeficiency virus (HIV)^6^), and variable surveillance systems across locations and time, inferring population-level measles immunity from a combination of vaccination coverage and case notifications can be challenging^7^.

Alternatively, serosurveys can provide a snapshot of immunity gaps that remain in a community by determining population-level prevalence of IgG antibody levels above specific thresholds that suggest clinical protection against disease.

As such, seroprevalence data can be used as tools to guide decisions to and strengthen immunization programs, as inputs to dynamic models of disease transmission, and additionally to provide insights into vaccine field effectiveness and assessment of case ascertainment rates^7,8^. The interpretation of seroprevalence data is complicated, however, because of the potential for bias. Some of this bias can be due to inadequate sensitivity of laboratory assays^9^ and/or specimen types^10^ used for measuring antibody levels. Additionally, bias from assay procedures can be suspected when protocols or commercial details are not reported or if no quality control was performed. Furthermore, population-based surveys have the potential for additional bias to be introduced in the selection of participants or from lack of representativeness of the selected sample from the community.

Beyond understanding the selection processes and laboratory assays used, it is critical to also consider how results of the serosurveys are reported. Considerations include what threshold of antibody titer was used as a correlate of clinical protection and how some tests report indeterminate results. In order to responsibly use and accurately interpret seroprevalence data for decision making or for modelling exercises, these issues need to be transparently acknowledged and discussed.

A more in-depth understanding of available seroprevalence data across locations and time, as well as the related implications, is critical for using these historic data to calibrate models used to inform decision making for immunization program strengthening, especially in low- and middle-income countries (LMICs) that face the highest ongoing measles burden. To fill these gaps, we first conducted a systematic review of literature reporting measles seroprevalence data published through 2021 and extracted information on key study and assay information.

Then, we developed a pilot bias assessment tool to assess the risk of bias in each study across the following categories: study selection of participants, measurement tool and classification of immunity, and results reporting.

## Methods

### Search strategy and selection criteria

This study follows PRISMA guidelines (Supplementary Tables 1-2) and was registered with PROSPERO (CRD42022326075). We performed a systematic review of published literature in any language containing information on population-level measles seroprevalence in LMICs. We searched PubMed on March 9, 2022 for primary data published through December 31, 2021 using the following search string:

(((Measles) AND (seroprevalence OR sero-prevalence OR seropositive OR sero-positive OR seronegative OR sero-negative OR seroepidemiology OR sero-epidemiology OR seroprofile OR seroimmunity OR sero-immunity)) OR (“Measles/epidemiology”[MeSH] AND (antibod* OR serolog*))) AND (“1900”[Date - Publication] : “2021”[Date - Publication])

One individual (ANS) screened titles and abstracts for each study in the search results. For relevant studies, one of multiple individuals (ANS, HF, IP) reviewed the full-text of each to determine their inclusion or exclusion. We included studies that contained original data on measles antibody prevalence and excluded studies if they only contained data from high-income locations (as based on WorldBank 2021 income classifications^11^), did not contain data on measles IgG antibody, were based on non-original data or from non-human subjects, contained only results from laboratory assay development or clinical trials (including studies only containing information on vaccinated persons), studied a target population of only health-care workers or active measles cases, or were a review, abstract, letter, editorial or brief report.

Following full text review, for each study that met our inclusion and exclusion criteria, we extracted the following data: study setting, study design and type (including information on planned, achieved (i.e., how many persons were reached via sampling), and reported (i.e., how many persons were represented in final study metrics) sample sizes), population demographics (including income and representativeness), type of specimen collected, serologic assay details (including type, name, and inclusion of a reference preparation), antibody threshold used for seropositivity and/or seroprotection (if relevant), and measures of proportion seropositive, seronegative, or indeterminate with accompanying uncertainty. We extracted data into a Microsoft Excel workbook and for seroprevalence measure, we recorded the most granular levels for relevant strata (i.e., by age, vaccination status, infection history, etc.) presented in each study.

### Bias assessment

Following extraction of all available data, we developed a comprehensive bias assessment tool and applied the tool to characterize the level of bias across each study. Our tool, modified from the ROBINS-I tool^12^, considers bias across the following categories, with associated indicators: study selection of participants, measurement tool and classification of immunity, and reporting of results (Supplementary Figures 1-3). We classified the level of bias across each category to be either low, moderate, severe, or critical. We then finally assessed the overall level of bias as low, moderate, severe, or critical for each study by taking the mean score of the category-specific classifications.

To assess bias among study selection of participants, we considered whether the study design used a random process for sample selection, if a study relied on a convenience sample, was restricted only to a subset of the population (e.g., only included pregnant women or cancer survivors), and reporting of planned, achieved, specimen, and final sample sizes. To assess the level of bias among the measurement tool and classification of immunity, we considered whether assay protocol, name, or references were provided, if internal or external validation or quality control was performed, and if there were other known factors known to decrease sensitivity or specificity. These factors included using oral fluid as specimens^13^, using a hemagglutination inhibition (HI/HAI) assay^13^, or using the Whittaker enzyme-linked immunosorbent assay (ELISA)^14^. Last, for bias among reporting of results, we considered whether a known threshold was used for determining protective titer levels, including metrics of uncertainty with seroprevalence estimates, and, if an enzyme immunoassay (EIA) or ELISA was used, whether and how equivocal results were handled and reported.

We characterized the overall level of bias in each study using the following criteria. For each category of bias studies were given a numeric score: low bias was assigned a score of 1, moderate a score of 2, severe a score of 3, and critical a score of 4. We took the mean of scores across all three categories. Studies with a mean score below 1.5 were characterized to have low overall bias, between 1.5 and 2.5 to have moderate overall bias, between 2.5 and 3 to have severe bias, and more than three to have critical bias.

We converted all metrics reported to proportion seropositive and then used R version 5.4.0 to compute summary metrics and make figures. For studies reporting seropositive and indeterminate/equivocal results independently, we did not include indeterminate results in the numerator of our overall seroprevalence calculation. We compared data availability by decade and bias level. We additionally investigated bias levels across time and region and assessed bias levels across locations with higher and lower first-dose measles-containing vaccine (MCV1) coverage^15^ and higher and lower estimated annual measles incidence^16^ in the year from which study data was collected.

### Role of the funding source

The Bill & Melinda Gates Foundation, Gavi, the Vaccine Alliance and the US National Institutes of Health had no role in study design, data collection, data analysis, data interpretation, or the writing of the report. All authors had access to the data and the corresponding author had final responsibility to decide to submit for publication.

## Results

### Systematic review

From our search, we identified 2032 studies for screening (Figure 1). Following screening, we excluded 1116 studies that did not meet our search criteria. For the remaining 916 studies, we assessed the full-text articles for inclusion. We identified 221 studies for inclusion and extracted information on measles seroprevalence, study design, and seroassay (link to zenodo file once uploaded). Studies were published between 1962 to 2021, including seroprevalence surveys conducted between 1953 and 2019.

**Figure 1.**
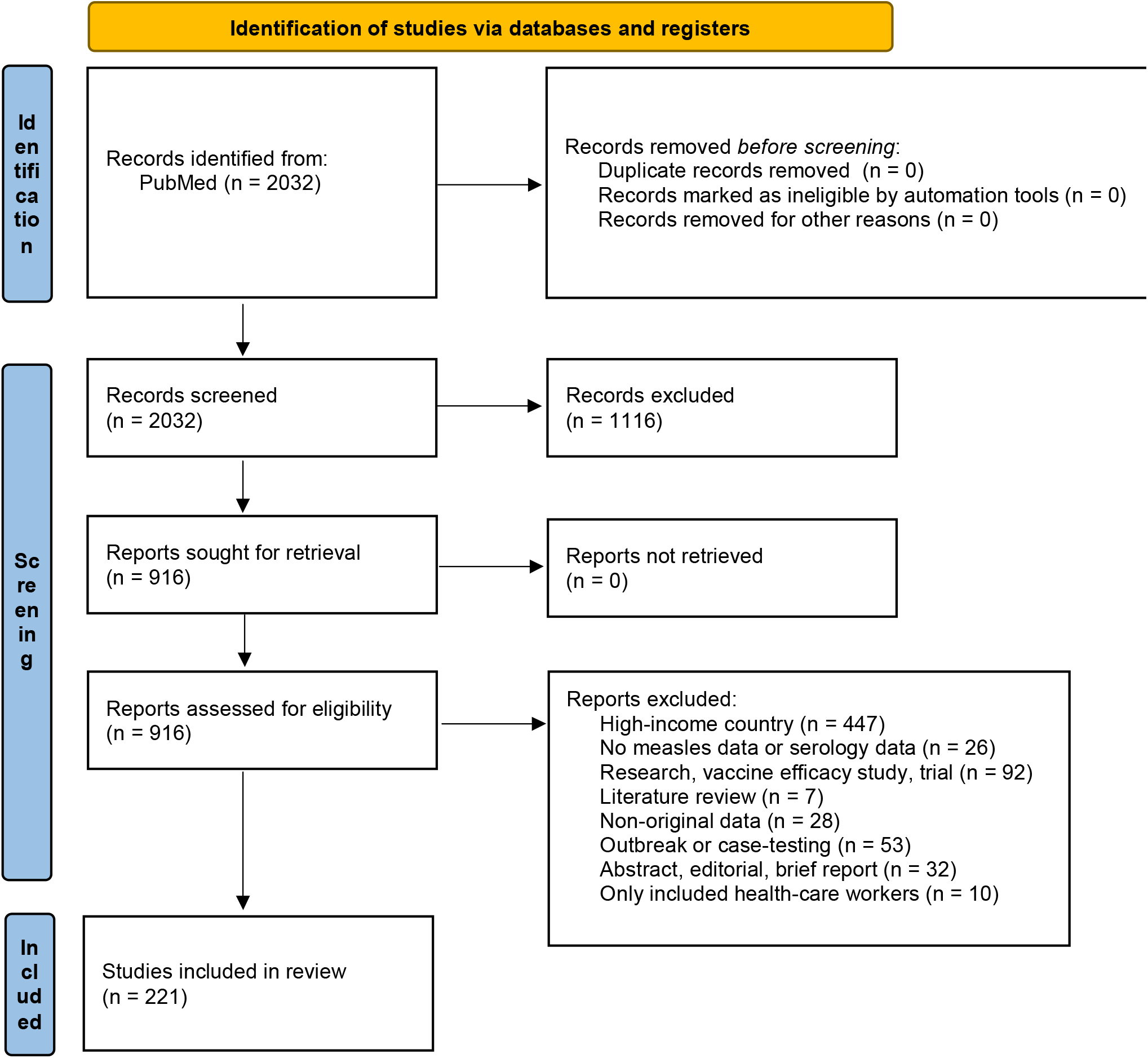
PRISMA diagram.

Among 182,789 persons sampled across all studies, age groups, and years, the crude mean measles seroprevalence was 78.00% (SD: 19.29%) and median seroprevalence was 84.00% (IQR: 72.75 – 91.66%).

Across regions of the World Health Organization (WHO), there were 43 studies containing data from the African Region, 47 from the Eastern Mediterranean Region, 35 from the European Region, 25 from the Region of the Americas, 20 from the South-East Asia Region, and 73 from the Western Pacific Region (Figure 2). There were 24 studies that represented data collected before 1980, 32 studies from 1980 to 1989, 29 studies from 1990 to 1999, 55 studies from 2000 to 2009, and 83 studies from 2010 to 2019. 178 studies (80.5%) contained age stratified results across 531 unique age ranges.

**Figure 2.**
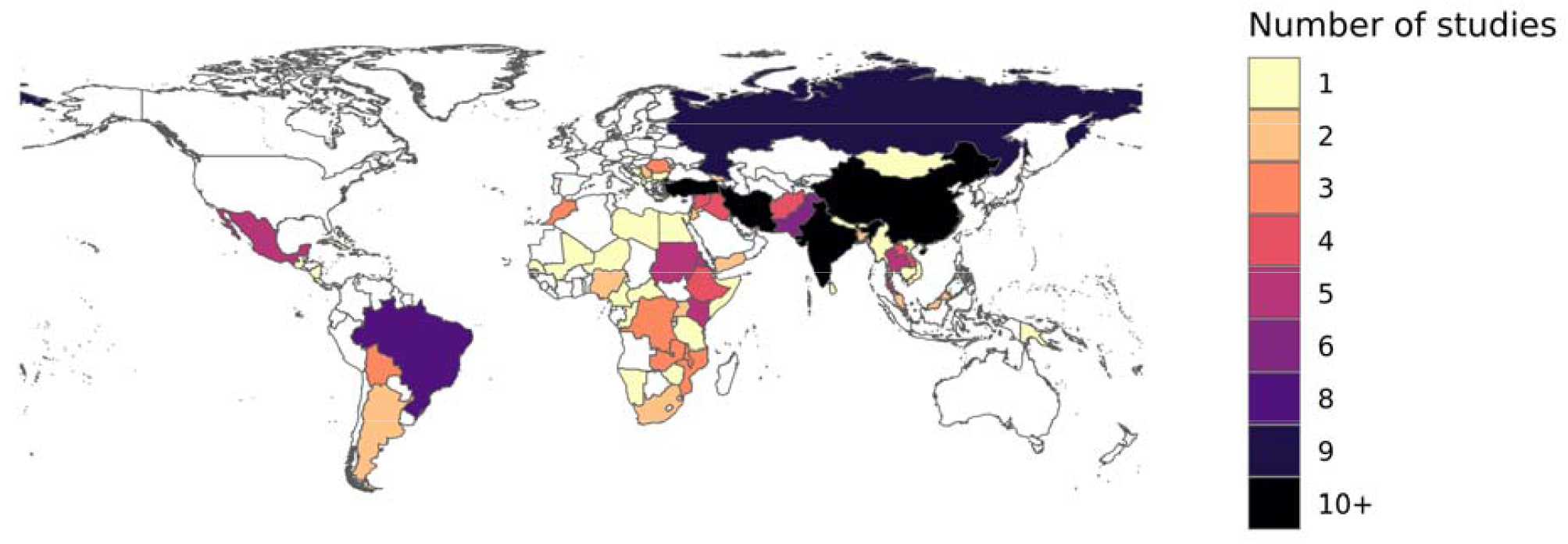
Number of serosurveys with data included per country. Map of number of studies per country with available data identified by systematic review.

### Bias assessment

Table 1 shows results of our bias assessment for each included study. For overall bias, we classified bias as low in 12 (5.4%) studies moderate in 129 (58.3%), severe in 58 (26.2%) and critical in 22 (10.0%). No studies had low or critical bias across all the categories of study selection of participants, measurement tool and classification of immunity, and reporting of results (Table 1).

**Table 1.** Overall and categorical bias classifications. Results of bias assessment in each of three categories (study selection of participants, measurement tool and classification of immunity, and reporting of results), and mean level of bias per study.

| Level of bias | Mean | Study selection of participants | Measurement tool and classification of immunity | Reporting of results |
| --- | --- | --- | --- | --- |
| Low | 12 | 15 | 19 | 20 |
| Moderate | 129 | 181 | 130 | 63 |
| Severe | 58 | 23 | 46 | 70 |
| Critical | 22 | 2 | 26 | 18 |

For study selection of participants, we identified 15 studies with low bias, 181 with moderate bias, 23 with severe bias, and 2 with critical bias. 81 studies used a random sample selection method. 117 studies with convenience samples used a restricted, non-representative sample (i.e., only among a specific subgroup of the population, such as persons living with HIV). 2 studies did not report the final sample size, and of the 81 samples that used a random sample selection method, 45 reported the planned sample size, and 15 additionally reported the planned, achieved, and specimen sample sizes.

In measurement assay and classification of immunity, we identified 19 studies with low bias, 130 with moderate bias, 46 with severe bias, and 26 with critical bias. Across the three categories of bias assessment, measurement assay and classification of immunity had the highest number of studies classified as having critical bias, largely due to absence of information on assay protocol details, commercial kit name or other appropriate citation describing the underlying methods.

195 studies provided details on the assay protocol or commercial kit name, and 25 studies conducted internal or external validation or quality control. 6 studies specified that samples were oral fluid specimens and 30 studies specified that samples collected were dried blood spots.

54 studies used an HI/HAI assay, 139 used an EIA or ELISA, 13 used a plaque reduction neutralization test (PRNT), 6 used a multiplex bead assay, and 11 used other or undescribed assay types. We noted changing temporal trends of types of seroassays used. While EIA, ELISA and PRNT assays were used in even distribution across all studies examined, there was no study published after 2001 that utilized an HI/HAI assay, and all studies using a multiplex immunofluorescent assay were conducted in 2013 or later.

We identified 20 studies with low bias, 63 with moderate bias, 70 with severe bias, and 18 with critical bias in reporting of results. 155 studies reported a threshold to define seroprevalence. Among the 139 studies that used an EIA or ELISA, 30 studies reported equivocal results separately or included with seropositivity results and 1 study excluded equivocal results and they were less than 5% of the overall sample. Finally, 59 studies reported metrics of seropositivity or seronegativity with any accompanying uncertainty.

### Seroprevalence trends

The crude median seroprevalence estimates from studies in the Western Pacific Region was 88.3% (IQR: 79.2 – 93.4%), in the Eastern Mediterranean Region was 87.2% (IQR: 81.3 – 93.2%), the European Region was 82.0% (IQR: 77.8 – 89.0%), in the Region of the Americas was 78.4% (IQR: 60.7 – 93.0%), in the African Region was 77.6% (IQR: 60.7 – 89.9%), and in the South-East Asia Region was 66.8% (IQR: 47.4 – 88.4%). Trends in seroprevalence and bias vary by decade (Figure 3). The median seroprevalence was lower in studies from 2010 to 2019 than those conducted before 1980 (i.e., the pre-vaccination era). Crude seroprevalence from studies conducted before 1980 was 90.5% (IQR: 67.8 – 93.3%), from 1980 to 1989 was 78.6% (IQR: 57.8 – 90.7%), from 1990 to 1999 was 88.3% (IQR: 60.7 – 92.6%), from 2000 to 2010 was 80.4% (IQR: 65.6 – 88.2%), and from 2010 to 2019 was 84.6% (IQR: 78.3 – 92.9%).

**Figure 3.**
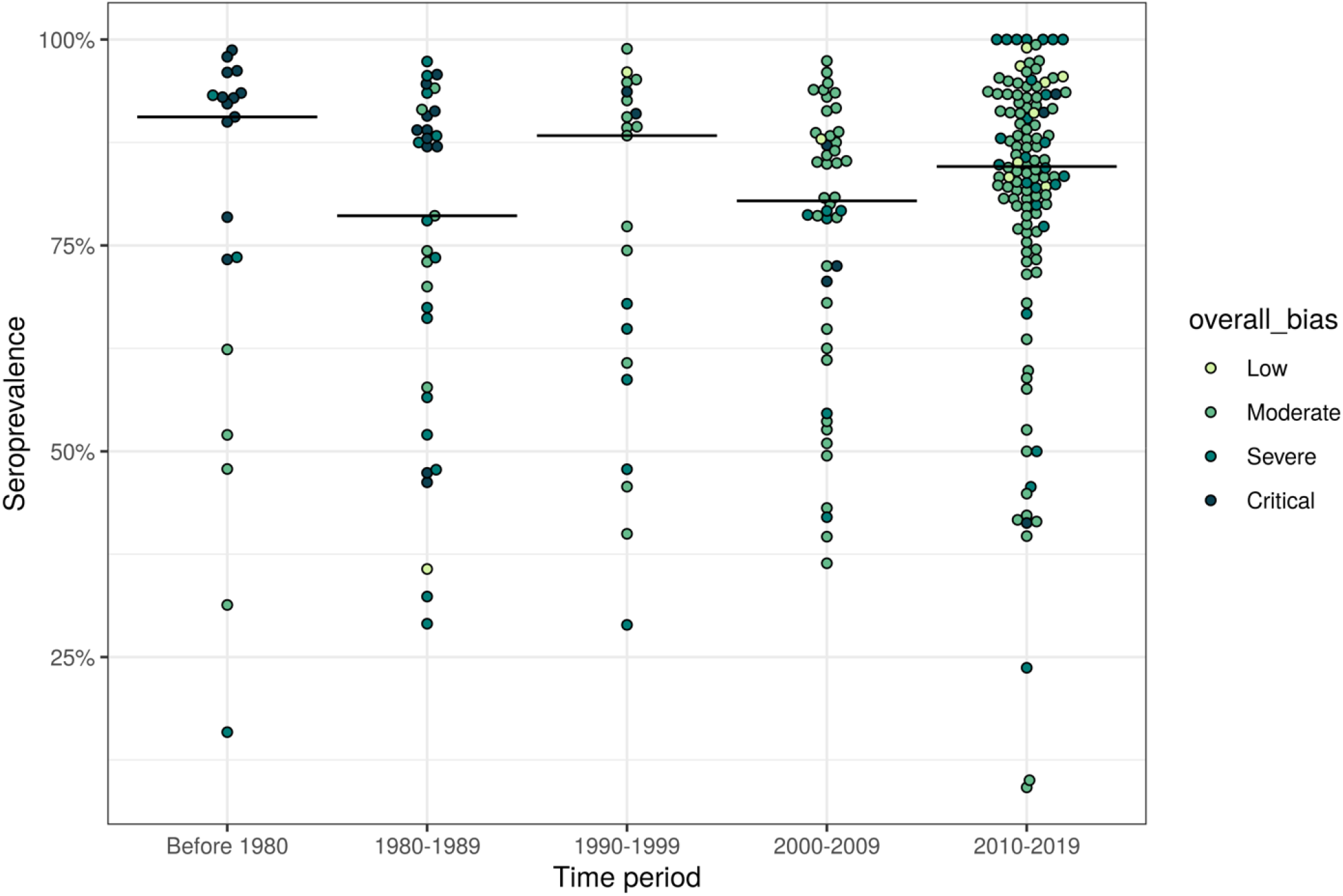
Measles seroprevalence by time period and overall bias level. Beeswarm plot of measles seroprevalence by time period. Each point represents one country-year of data per study and are colored by overall bias level. Black lines represent the median observation across each decade.

Among 31 country-years with studies containing critical bias, 23 (74%) occurred in earlier time periods (i.e., before 1980 and between 1980 and 1989). In the 159 country-years with studies containing low or moderate bias, 96 (60%) have occurred between 2010 and 2019.

We additionally compared the overall bias levels for each country-year of the studies to the MCV1 coverage and measles incidence from the same country-year (Figure 4). Generally, studies in countries and years in 1980 or later with lower MCV1 coverage and higher measles incidence had more bias compared to studies from countries and years with higher MCV1 coverage and lower measles incidence (p < 0.001, in proportional odds logistic regression models for both MCV1 coverage and incidence). Among 109 studies from countries and years with MCV1 coverage greater than 80%, 93 (85%) had low or moderate overall bias, and from the 58 studies from countries and years with MCV1 coverage of 80% or lower, 34 (58%) had low or moderate overall bias. A similar trend persisted across studies in countries and years with high incidence – 103 of 122 (84%) studies in countries and years with average annual reported measles incidence less than 5 per 1000 persons had low or moderate overall bias, and 24 of 49 (49%) of studies in countries with annual measles incidence of 5 per 1000 persons or greater had low or moderate overall bias.

**Figure 4.**
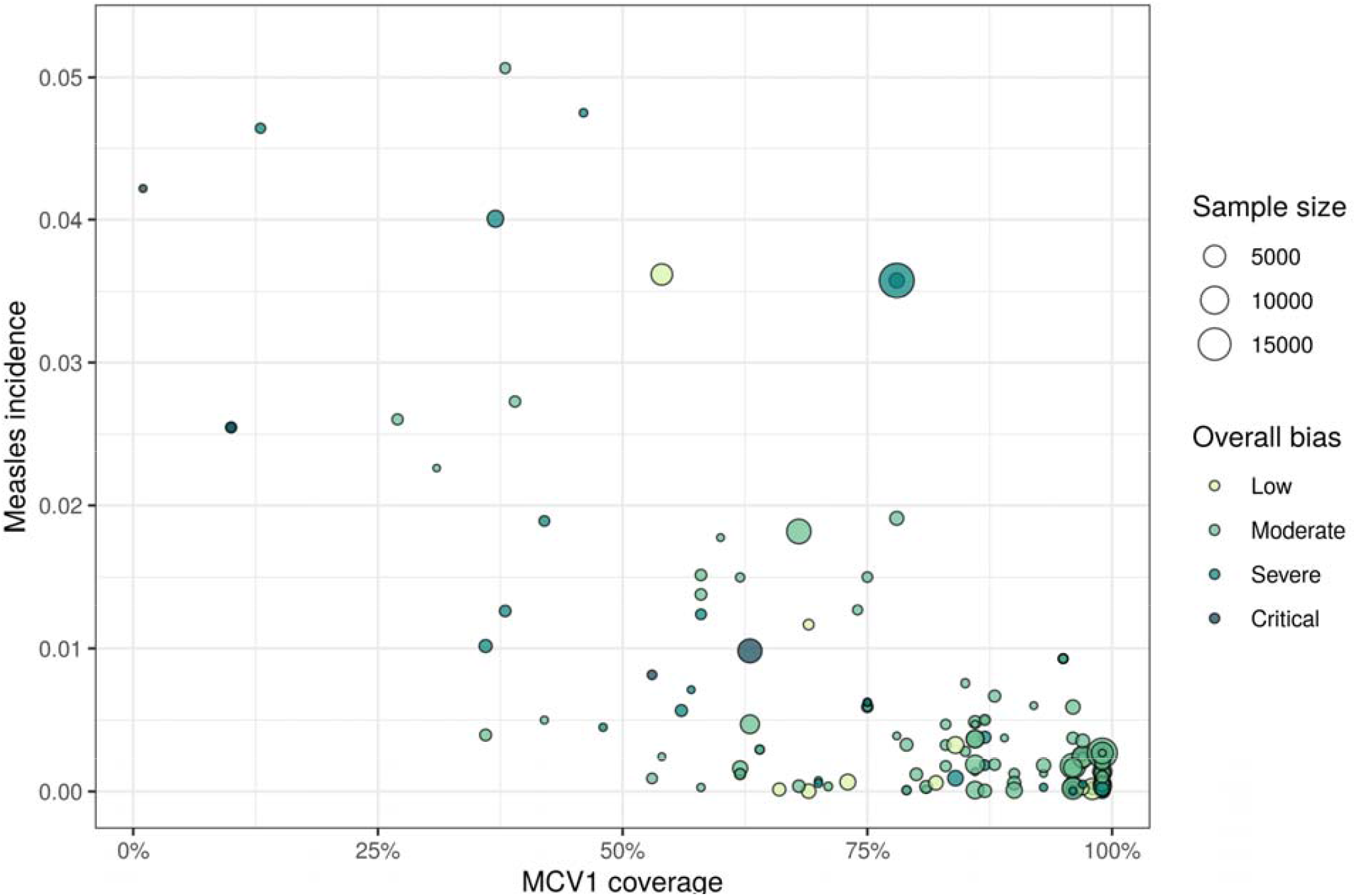
Overall bias level by MCV1 coverage and annual measles incidence. Each point represents each country-year represented across all studies, colored by overall bias level, by MCV1 coverage and annual estimated measles incidence.

## Discussion

To identify the scope of measles seroprevalence data, we conducted an updated systematic review of serosurveys to identify primary data sources and characterized underlying bias across these studies. The resulting data repository from our investigation along with information on factors related to underlying bias per study could contribute to analyses of measles dynamics among low- and middle-income countries. We identified serosurveys available in each decade, WHO region, and across a wide variety of ages, which could be useful when modelling location-, time-, and age-specific estimates of measles transmission and susceptibility. Despite this variation, there were locations for which very few or no serosurveys have been conducted – mainly in the African Region – which contribute to knowledge and data gaps to inform high-quality modelling and analyses.

Additionally, our study provides insight to issues to consider when designing and reporting a seroprevalence study to ensure that the highest quality surveys are conducted and that complete, accurate and transparent reports are generated. The number of available measles seroprevalence studies has increased in the last few decades compared to periods before the introduction of national measles vaccination programmes in LMICS. This trend provides the opportunity for researchers to examine the impact of vaccination programs on ongoing susceptibility within the population represented in each study. However, we found that locations with high annual measles incidence and lower MCV1 coverage tend to have not only less studies conducted, but also higher bias – this is understandable given that coverage tends to be lower in the most difficult settings such as remote and/or conflict-affected regions, where surveys are especially challenging to conduct. Research and programmatic teams planning seroprevalence studies, especially among persons living in these vulnerable communities, could use the framework presented in this study as a starting point to determining the feasibility and cost of conducting a high-quality seroprevalence survey and consider alternative ways to invest the funds (e.g., in strengthening ongoing surveillance of coverage and disease incidence).

More recently, there have been examples of high-quality serosurveys, such as a nationally representative survey in Zambia^17^, that have been conducted and used for informative modelling. Given the complexity, time, and expense of these surveys, it is worthwhile to make the most of high-quality surveys that are being conducted for different infections and funded through a variety of different programs. This serosurvey in Zambia, for example, leveraged residual sera from the Zambia Population-Based HIV Impact Assessment (ZAMPHIA) study^18^ originally collected to estimate HIV incidence and viral load. Applications of such data extend to innovative modelling efforts to estimate subnational and age-specific seroprevalence estimates as well as national level outbreak risk^17^. That study serves as an example of the potential to leverage other major population surveys and to use high quality seroprevalence estimates to inform evidence for decision making.

More studies had low or moderate bias compared to severe or critical bias among the categories of selection of study participants and measurement tool and classification of immunity. For the category of reporting of results, more studies had severe or critical bias levels than low or moderate bias levels. Overall, we found that less than 10% of studies had low overall bias, suggesting that the quality of conduct and reporting of seroprevalence studies has substantial potential for improvement.

While interpreting seroprevalence estimates identified by our review, it is essential to also consider the associated sensitivity and specificity of the seroassays used in studies along with the route of induced immunity (i.e., from vaccination or natural infection). For example, HI/HAI assays are often less sensitive than other types^13^. If HI/HAI assays are used in a population with mainly vaccine-induced immunity, seroprevalence results may be underestimated. However, since HI/HAI assays were historically used more frequently, during an era with less vaccine-derived immunity and subsequently higher natural immunity affording higher antibody levels, assay sensitivity might not be as important to consider. In our bias assessment in the category of measurement tool and classification of immunity, we defined factors that influence assay specificity and sensitivity as either (1) using an HI/HAI assay, (2) using the Whittaker commercial ELISA kit, or (3) using oral fluid samples. However, the utility of this specific contribution to our bias assessment might be subject to the specific study setting, vaccination program implementation and success, and underlying measles epidemiology.

Our study has several limitations. First, we were unable to fully synthesize results of our systematic review in a meta-analysis or other stratified analysis by age, location, or year. This was due to the differing study populations, regions, time periods, and age groups presented in studies identified in this review as well as the varying degrees of bias characterized to be present across studies. These results can serve as the basis for future models that synthesize the data while also accounting for underlying measles infection dynamics, vaccination coverage and population structures for each individual study setting, which was out of the scope of our analysis.

Secondly, we were constrained by the information reported in each publication. Without adequate reporting, we assumed the highest level of associated bias whenever appropriate. For example, if a study did not specifically note if they used an international reference preparation, we assumed they did not use one. This may have led us to classify studies as having higher bias in relevant categories than might have been the case if all available information had been included in the publication – it possible that some details were omitted to meet restrictions on word counts, for example. As such, there might be great utility in the widespread use of standardized reporting expectations for ongoing and future seroprevalence studies.

Next, we did not consider sample size in our assessment of bias. Since the impact of sample size on the reliability of point estimates from seroprevalence studies should be reflected in the provided uncertainty interval, we considered the inclusion of such in our bias assessment. We did not however further assess the implications of smaller or wide interval spans of if they were presented or whether point estimates or uncertainty intervals were adjusted or standardized for population demographics or other factors. Finally, there are likely additional sources of bias that are more difficult to ascertain objectively, such as potential issues with specimen storage and laboratory capacity, practices, and quality.

Our study strengthens the understanding of the availability and bias among measles seroprevalence studies in low- and middle-income countries by identifying primary sources of measles seroprevalence studies and conducting a bias assessment of the associated data. Our framework for assessing bias could provide a foundation for further work by relevant agencies and interested partners to develop a tool for use in planning and reporting future surveys. This work can be a vital tool to be used during modelling exercises, planning immunization-based interventions, and ultimately, to make informed decisions to reduce preventable measles morbidity and mortality.

## Data Availability

All data produced in the present study are available upon reasonable request to the authors.

## Contributions

ANS, MJ and JFM conceived and planned this study. ANS, FC, DR, MJ and JFM designed the bias assessment framework. ANS, HF and IP screened and extracted studies. ANS made tables and figures. ANS wrote the first draft of the manuscript and all authors contributed to subsequent revisions.

## Notes

### Competing Interest Statement

ANS, HF, JFM and MJ are funded by the Bill & Melinda Gates Foundation, Gavi, the Vaccine Alliance. ANS receives funding from the US National Institutes of Health. MJ receives additional funding from Wellcome Trust
UKRI, MRC, NIHR and US CDC. All other authors declare no competing interests.

### Funding Statement

This study was funded by the Bill & Melinda Gates Foundation, Gavi, the Vaccine Alliance, and the US National Institutes of Health.

